# Inspiratory Muscle Strength Training to Improve Cardiometabolic Health in Patients with Type 2 Diabetes Mellitus: Protocol for the Diabetes Inspiratory Training (DIT) Clinical Trial

**DOI:** 10.1101/2023.10.27.23297688

**Authors:** Baylee L. Reed, Dallin Tavoian, E. Fiona Bailey, Janet L. Funk, Dawn K. Coletta

## Abstract

Type 2 diabetes mellitus (T2DM) is a complex, chronic metabolic disease that carries with it a high prevalence of comorbid conditions, making T2DM one of the leading causes of death in the U.S. Traditional lifestyle interventions (e.g., diet, exercise) can counter some adverse effects of T2DM; however, participation in these activities is low with reasons ranging from physical discomfort to lack of time. Thus, there is a critical need to develop novel management strategies that effectively reduce cardiometabolic disease risk and address barriers to adherence. High-resistance inspiratory muscle strength training (IMST) is a time-efficient and simple breathing exercise that significantly reduces systolic and diastolic BP and improves vascular endothelial function in adults with above-normal blood pressure. Herein, we describe the study protocol for a randomized clinical trial to determine the effects of a 6 week IMST regimen on glycemic control and insulin sensitivity in adults with T2DM. Our primary outcome measures include fasting plasma glucose, fasting serum insulin, and insulin sensitivity utilizing homeostatic model assessment for insulin resistance (HOMA-IR). Secondary outcome measures include casual (resting) systolic BP and endothelial-dependent dilation. Further, we will collect plasma for exploratory proteomic analyses. This trial seeks to establish the cardiometabolic effects of 6 weeks of high-resistance IMST in patients with T2DM.

## Introduction

Type 2 diabetes mellitus (T2DM) is at epidemic proportions in the United States, affecting 37.3 million people (or 11.3% of the population) [1]. In 2017, the annual economic cost was $327 billion, making it one of the most expensive chronic conditions in the U.S. [2]. T2DM is a chronic, obesity-associated metabolic disorder characterized by glucose dysregulation, insulin resistance, and beta-cell defects [3,4]. Adults with T2DM are likely to develop vascular endothelial dysfunction and vascular inflammation, increasing their risk of cardiovascular disease (CVD) and the occurrence of cardiac events [5]. First-line T2DM treatments include lifestyle modifications, such as dietary changes and exercise [6], the benefits of which are well established in T2DM patients [7,8]. However, adherence to conventional exercise is low, as only 41% of U.S. adults with T2DM meet current aerobic exercise guidelines, and only 12% meet resistance exercise guidelines, compared to the general population, with participation rates at 52% and 21%, respectively [9]. This is due in part to barriers such as physical discomfort and lack of time [10,11]; thus, new regimens designed to overcome these barriers are needed.

Recently, a novel and time-efficient respiratory exercise called Inspiratory Muscle Strength Training (IMST) was developed [12]. IMST is distinct from other traditional forms of exercise due to its abbreviated training format (i.e., 5 minutes daily) and is performed using a hand-held device while seated or standing [12]. With just six weeks of training (5 days/week), high resistance-IMST has been shown to lower systolic blood pressure (systolic BP) by ∼9 mmHg in normotensive and hypertensive adults [13]. Furthermore, it has improved endothelial-dependent dilation (EDD) by 45% in older adults with impaired endothelial function [14]. These vascular effects of IMST are believed to reduce the risks of CVD, the number one cause of death in people with T2DM [15]. IMST is safe and well tolerated, with adherence rates >90% in diverse populations [14,16], and thus presents a manageable introductory or adjunctive program for improving cardiometabolic health in T2DM patients who have difficulty maintaining a traditional exercise program. However, the effects of IMST on glycemic control and insulin sensitivity, as well as systolic BP and EDD, are unknown in patients with T2DM.

Vascular endothelial function and metabolic function are closely linked [17]. The vascular endothelium produces nitric oxide (NO), which is released in response to increased arterial wall shear stress (i.e., increased blood flow) [18]. Among its various functions, NO enhances glucose uptake into cells and improves insulin sensitivity [19]. T2DM is associated with impairments in endothelial function, including reduced NO production and increased vascular inflammation [20]. High resistance-IMST is a potentially effective tool to combat T2DM-associated endothelial dysfunction, as it has been shown to increase NO bioavailability and reduce oxidative stress [14] — key adaptations that could underly the improved metabolic health. The latter is especially significant given the link to metabolic syndrome, which encompasses insulin resistance, impaired glucose metabolism, and hypertension [21] and, therefore, heightens the risks for cardiac events or stroke [21].

The potential for IMST to elicit cardiometabolic adaptations in patients with T2DM warrants assessment. Accordingly, we outline a plan to interrogate the effects of 6 weeks of high-resistance IMST on glycemia (fasting plasma glucose), insulin sensitivity/resistance (fasting serum insulin and Homeostasis Model Assessment [HOMA-IR; ratio of fasting insulin/glucose]), causal (resting) BP, and NO-mediated EDD in T2DM patients. Participants will be randomized into either high-resistance (experimental) or low-resistance (control) groups and complete IMST at home 5 days/week for 6 weeks, with each session lasting ∼5 minutes [22].

We will study T2DM patients before and after 6 weeks of high-resistance IMST to test the hypotheses that (1) fasting plasma glucose will decrease, and insulin sensitivity will improve, (2) Casual (resting) systolic BP will decrease, and (3) high-resistance IMST will improve EDD resulting in clinically-meaningful improvements (i.e., >1% unit change) [23]. Lastly, we will consent and collect DNA for banking for future studies and plasma to perform quantitative proteomics to evaluate novel protein expression changes pre-versus post-IMST. Our goal from the exploratory proteomic analyses is to identify mechanisms that underlie the training.

## Materials and Methods

### Study Design

The Diabetes Inspiratory Training (DIT) study is a randomized, sham-controlled, exploratory clinical trial examining the effects of IMST in 24 adults with T2DM. This is a 6 week intervention study design. An outline of the study is shown in Figure 1.

**Figure 1.**
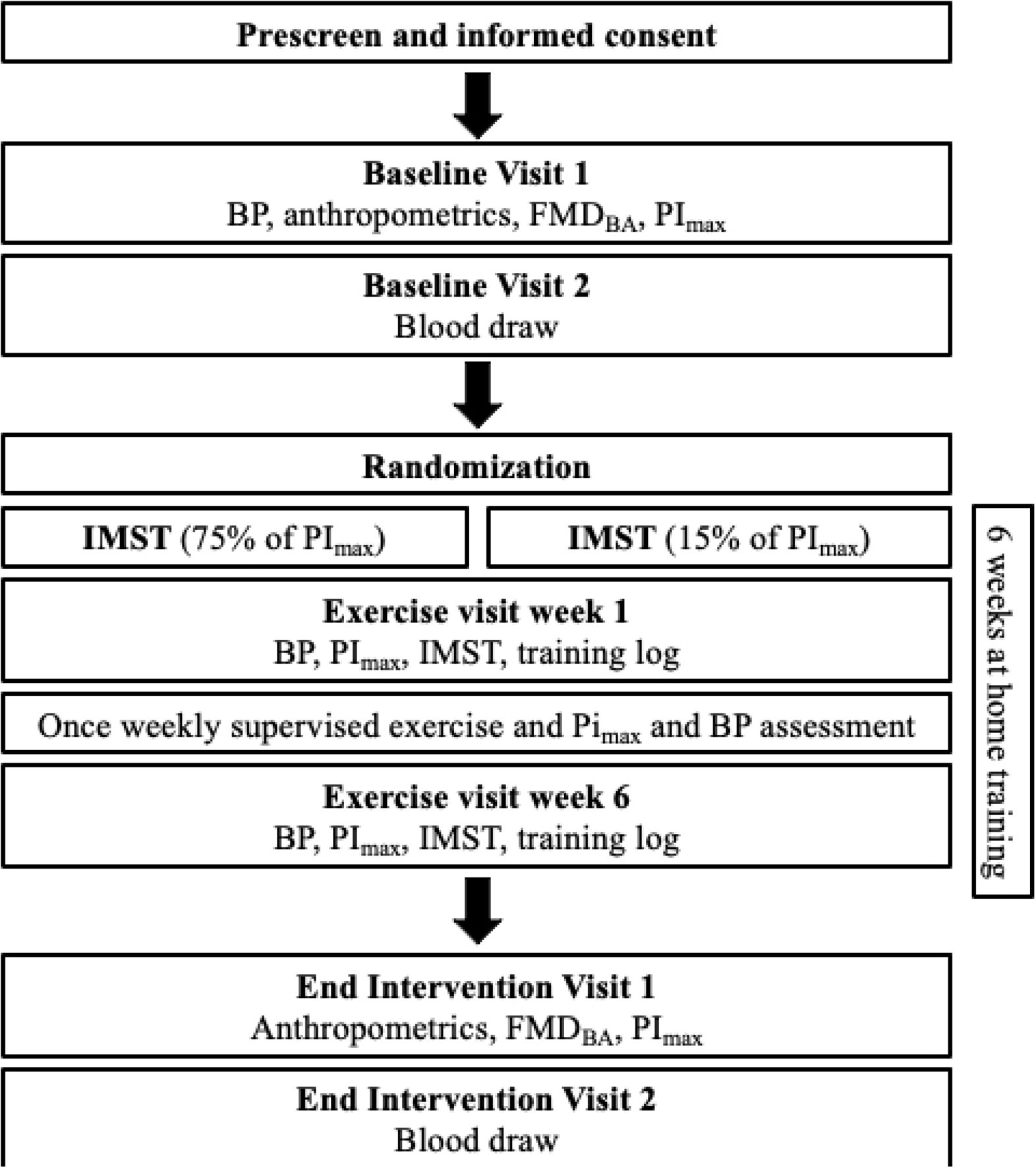
Study Flow for the DIT Study

Participants will perform 5 sets of 6 breaths per day, 5 days per week, at either high relative resistance (75% of maximal inspiratory pressure in cmH_2_O, (PI_max_)) or low relative resistance (15% of PI_max_) [22].

### Participants and Interventions

#### Study Setting and Study Population

The DIT Study will be conducted at the Clinical and Translational Sciences (CATS) Research Center at the University of Arizona. Participants will be pre-screened over the phone to determine their eligibility using the inclusion and exclusion criteria (Table 1).

**Table 1.** Inclusion and Exclusion Criteria for DIT Study.

| Inclusions | Exclusions |
| --- | --- |
| 18+ years old | Current smoker (tobacco / cannabis) |
| Diagnosed with type 2 diabetes by physician | Uncontrolled medical condition (e.g., cancer) |
| Fasting plasma glucose levels $\geq 126$ mg/dl and $\leq 240$ mg/dl | Myocardial infarction or stroke within the previous 12 months |
| SBP between 120-169 mmHg | Performs regular aerobic exercise ($>4$ bouts/week) |
| Stable dose of medication (3 months on same dose) | DBP $>100$ or $<60$ mmHg |
| Weight stable in the prior 3 months ($< 3$ kg weight change) and willing to remain weight stable throughout the study | SBP $<120$ or $\geq 170$ mmHg |
| Absence of unstable clinical disease as determined by medical history | Medications that, in the opinion of the study physician or nurse practitioner, may impact the outcomes of the study (e.g., steroids) |
|  | Cheyne-Stokes Respiration |
|  | History of perforated eardrum |
|  | History of glaucoma or retinopathy |
|  | Pregnant, breastfeeding or trying to become pregnant (self-reported) |

SBP= systolic blood pressure, DBP = diastolic blood pressure, mmHg = millimeters of mercury, mg/dl = milligrams per deciliter, kg = kilograms, kg/m<sup>2</sup> = kilograms per meter squared

#### Eligibility Criteria

Participants will be eligible for this study if they 1) are 18 years of age or older, 2) have been previously diagnosed with T2DM by a physician, 3) have a systolic BP between 120-169 mmHg, 4) are on a stable dose of medication for at least 3 months, 5) do not have an unstable clinical disease, and 6) do not meet the exclusion criteria (Table 1).

#### Interventions

All training will be completed using the POWERbreathe^TM^ K3 trainer (POWERbreathe International Ltd., Warwickshire, U.K.). This is a handheld pressure-threshold device with a computerized threshold sensor. Each participant will be provided their own device to perform IMST at home, 5 days/week for 6 weeks. They will receive in-person verbal instruction on the training protocol and K3 operation from the Research Technician at the start of the study.

The Research Technician will monitor one training session weekly in the CATS facility; the remaining 4 sessions will be completed unsupervised at home. During each visit to the CATS facility, the Research Technician will determine the participant’s PImax and transfer the saved training data from the K3 device to ensure exercises are being completed at home. The PImax will be determined by taking the average of 3 measurements. The Research Technician will then adjust the training resistance as needed to ensure participants are training at the prescribed intensity (i.e., either 15% or 75% of PImax). The participant will then perform a supervised training session. All training sessions will be recorded in the device memory card, and participants also will be required to complete a weekly training log.

### Outcomes

#### Primary Outcomes

##### Fasting Plasma Glucose and Fasting Serum Insulin

Participants will report to the CATS facility at the University of Arizona following a 12-hour fast for both the baseline and post-intervention visits. Up to 5 mL of blood will be drawn from the antecubital vein and sent to Sonora Quest for screening laboratory tests, lipid measures, and metabolic panels, including fasting plasma glucose and fasting serum insulin.

##### Insulin Sensitivity

Insulin sensitivity will be calculated using the Homeostatic Model assessment for insulin resistance (HOMA-IR), which is validated method of measuring insulin sensitivity [24]. The equation for HOMA-IR is:

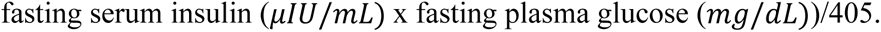

#### Secondary Outcomes

##### Casual (Resting) Blood Pressure

We will measure casual (resting) blood pressure per the American College of Cardiology (ACC) and the American Heart Association (AHA) guidelines [22] with an automated oscillometric sphygmomanometer (SunTech CT40, SunTech Medical). Briefly, participants will be asked to sit quietly with both feet flat on the ground, backs supported, and with their arms resting at heart level [22]. Three measures will be performed after a 5 minute quiet rest period with 1 minute of recovery between each measure. The average systolic and diastolic blood pressure will be recorded pre- and post-intervention.

##### Endothelial Dependent Dilation

Endothelial Dependent Dilation (EDD) will be assessed via brachial artery flow-mediated dilation (FMD) using high-resolution ultrasonography (Canon Xario 200G), as previously described [25]. Participants will be asked to avoid exercise, caffeine, and alcohol for 24 hours and food for at least 5 hours prior to their visit. FMD will be assessed by measuring the brachial artery diameter and blood velocity at baseline and for 3 minutes following reactive hyperemia, which stimulates NO release. Reactive hyperemia is induced by 5 minutes of forearm blood flow occlusion with a cuff placed on the upper forearm and inflated at least 50 mmHg above systolic BP [22,26]. Brachial artery diameter and blood velocity will be analyzed offline using commercially available software (Brachial Analyzer, Medical Imaging Applications LLC, Coralville, IA, USA) [22] and expressed as absolute (mm) and percent change in arterial diameter from baseline (pre-cuff inflation diameter) to post-intervention following the 6 weeks of IMST.

#### Exploratory Outcomes

##### Proteomic Analysis

Blood will be collected into purple K2-EDTA vacutainers and immediately placed on ice, then centrifuged at 3,000 rpm at 4°C within 10 minutes of blood collection. Separated plasma will be removed and frozen at -80°C in cryotubes until analyzed using high-performance liquid chromatograph-electrospray ionization-MS/MS (LC-MS) [27]. Briefly, the extracted plasma proteins will be subjected to subsequent in-solution digestion using trypsin and Lys-C to be analyzed with tandem mass spectrometry [27,28]. Lastly, quantitative proteomics will be performed using extracted ion abundance, including statistical analysis via Progenesis [29]. The resulting quantitative proteomic data sets will be analyzed using DAVID for gene ontology and pathway enrichment analysis [30,31].

##### DNA banking

Blood will be collected from the antecubital vein of the arm directly into PAXgene DNA collection tubes, as per the manufacturer’s instructions. Briefly, these tubes contain an additive reagent that stabilizes the blood. The tubes will sit at room temperature for 2 hours and then be stored in the -20 freezer until processed. The PAXgene DNA processing kit will be used to isolate the DNA. Once DNA is extracted, it will be stored and banked for future studies. Participants will be required to provide their consent for banking of their de-identified DNA/plasma samples.

#### Participant timeline

The timeline for participation in the study will be 7-9 weeks, as shown in Figure 1. A summary of the visits for the DIT study participants is shown in Table 2.

**Table 2.** Summary of the Visits for the DIT Study.

|  | Baseline |  | Training | End-Intervention |  |
| --- | --- | --- | --- | --- | --- |
|  | Visit 1 | Visit 2 | Week 1-6 | Visit 1 | Visit 2 |
| **Screening** |  |  |  |  |  |
| Informed Consent | X |  |  |  |  |
| Demographics | X |  |  |  |  |
| Medical History | X |  |  |  |  |
| Inclusion/Exclusion | X |  |  |  |  |
| Hip/waist/neck circumference | X |  |  | X |  |
| **Functional Assessments** |  |  |  |  |  |
| Casual BP | X |  |  | X |  |
| PI<sub>max</sub> | X |  | X | X |  |
| Spirometry | X |  |  | X |  |
| **Blood Draw** |  |  |  |  |  |
| Metabolic panel |  | X |  |  | X |
| Plasma / DNA |  | X |  |  | X |
| **Arterial Assessments** |  |  |  |  |  |
| FMD<sub>BA</sub> | X |  |  | X |  |
| **At-home Testing** |  |  |  |  |  |
| Sleep/exercise diary |  |  | X |  |  |
| **Exercise sessions** |  |  |  |  |  |
| Daily exercise |  |  | X |  |  |
| Once-weekly supervised |  |  | X |  |  |
| Weekly text/email |  |  | X |  |  |
| **Randomization** | X |  |  |  |  |

BP = Blood Pressure, FMD<sub>BA</sub> = Flow-mediated dilation of brachial artery, PI<sub>max</sub> = Maximal inspiratory mouth pressure

Participants will sign a written informed consent with a member of the research team at the CATS facility. Following informed consent, participants will complete all baseline assessments in two in-person visits to the CATS facility within a 14-day window. Participants will begin the 6 week intervention ≤ 14 days after baseline assessments are completed. All assessments will be repeated within 14 days after completing the 6 week training program. Participants will continue to perform IMST 5 days/week until all post-assessments are completed.

#### Power analysis and sample size

A minimum of 16 and a maximum of 24 participants will be enrolled and randomized into groups. To our knowledge, the effect of IMST, specifically 75% resistance, on fasting glucose and/or insulin sensitivity in any population has not previously been reported, nor have effects of IMST on systolic BP and EDD in T2DM been specifically ascertained. Thus, for our power analysis, we estimated a modest effect size of 0.40 with alpha set at 0.05 using repeated measures ANOVA within-between framework. A sample size of 16-24 will have 85-96% power for any outcome with an effect size ≥ 0.40.

#### Recruitment

Recruitment will be via word of mouth, advertisements placed in area newspapers, social media, and flyers posted around the University of Arizona and to the surrounding local community in Tucson. Interested individuals will be directed to the study website, where they will be able to complete a questionnaire to determine their eligibility. Individuals who do not meet the inclusion criteria will be informed of their ineligibility. Candidates who meet eligibility will be contacted for a study overview session.

After the study overview, written informed consent will obtained in person from each participant before the start of any study-related procedures. Ethical Approval for this study has been obtained from the University of Arizona Institutional Review Board (Protocol 00002239).

### Assignment of Interventions

#### Sequence generation

The randomization sequence was created using computer-generated random numbers at a 1:1 ratio in blocks of four. Male and female participants will be randomized using separate randomization tables.

#### Allocation concealment mechanism

Group allocation will be stored in an Excel file that is not available to the Research Technician.

#### Implementation

Once the Research Technician has completed all enrollment activities for a participant (i.e., a participant has met the inclusion criteria and completed baseline assessments), the Principal Investigator (PI) will inform the research technician of the participant’s allocation group.

#### Blinding

Due to the nature of the study, the participants are blinded to the intervention.

### Data Collection, Management, and Analysis

#### Data management

Data will be collected with paper data collection forms and entered into a Microsoft Excel sheet within 48 hours of data collection. At the end of the study the Excel sheet will be rechecked against the paper originals and any inconsistencies will be noted and discussed between the PI and Research Technician in charge of data entry.

#### Statistical plan

Data will be analyzed with a repeated measures ANOVA test and Sidak post hoc testing using SPSS version 28.0. Group-by-sex interactions will be investigated, and effect sizes with confidence intervals will be reported in addition to p-values. All tests will be two-sided with alpha set at 0.05.

### Monitoring

#### Data monitoring

The intervention is low-risk and does not require a data monitoring committee. The research team will meet with the study physician at regular intervals to track study progress and discuss any potential safety issues. No interim analyses will be performed.

#### Harms

An adverse event (AE) is any harmful and unintended reaction during the course of the study that may be related or unrelated to the intervention. All AEs occurring between a participant signing the informed consent and completing post-intervention assessments will be reported to the study physician.

### Anticipated Results

#### Primary Hypothesis

Six weeks of high-resistance IMST will lower fasting plasma glucose and improve insulin sensitivity.

#### Other Hypotheses

Six weeks of high-resistance IMST will:

1. Lower causal (resting) systolic BP
2. Improve EDD

## Discussion

Regular exercise is one of the most commonly prescribed non-pharmacological interventions for T2DM management and yields improvements in glycemic control and insulin action [7,8]. However, aerobic exercise is physically strenuous and time-consuming [10,11], and less than half of T2DM adults participate in exercise on a regular basis. In contrast, IMST is a novel form of high intensity training that can be performed whether sitting or standing, requires only 5 minutes per day, and rapidly improves blood pressure, endothelial vascular function, and vascular resistance among hypertensive adults [13,14,16,32]. Whether IMST can also affect changes in fasting blood glucose or insulin sensitivity is of critical interest and important for adults with T2DM, along with establishing if these blood pressure lowering effects and increased EDD are also seen in this population following IMST.

A study by Corrêa et al. studied the acute effects of IMST on glucose variability and showed significant improvements in glucose immediately following the training [33]. Additionally, another study, which was for 12 weeks at a lower resistance of 30% revealed no significant changes in blood glucose levels [34]. The discrepancies across these findings are likely due to the populations studied, the timeframe of the training, and the resistance used. To our knowledge, there have been no investigations that have reported the effects of chronic IMST training at a resistance of 75% on glycemic control and insulin sensitivity in T2DM.

High-resistance IMST could yield long-term beneficial effects in T2DM patients, as it has in other populations [12,14,16,22,32]. We anticipate a reduction in fasting plasma glucose and improved insulin sensitivity while seeing similar established physiological benefits such as reduced systolic BP and improved endothelial function.

## Ethics and Dissemination

### Research ethics approval

This study has been approved by the University of Arizona Institutional Review Board (Approval Number: 00002239).

### Protocol amendments

Any modifications to the protocol that may impact the conduct of the study will first be decided by the PI and approved by the University of Arizona IRB prior to any implementation. Administrative changes of the protocol are considered minor corrections that have no impact on the way the study is to be conducted. These changes will be agreed upon by the PI and documented.

### Consent

This study will be thoroughly explained to each participant in person where subjects will have the opportunity to ask questions. Once the member of the research team believes the participant understands the study requirements, they will be directed to read and sign the informed consent document.

### Confidentiality

Identity of the participants will be protected by assigning each a code (i.e., a 3-digit number) and any experimental data collected from these subjects will be recorded under that number. Any identifiable personal information will be kept in a password-protected digital file and/or in a locked cabinet. Only the PI, Co-I and Research Technician will have access to the information.

### Access to data

The PI, Co-I and Research Technician will have access to the final trial dataset. Other project team members will be provided de-identified data for their analysis.

### Ancillary and post-trial care

There are no provisions for ancillary or post-trial care.

### Dissemination policy

Primary outcome papers will be approved by the PI prior to journal submission. Every attempt will be made to release study results to the general public soon after study completion. Interim and final reports may also be presented at various local, regional, and international conferences, with approval from the PI. Eligibility for authorship include (1) substantial contribution to study conception and design AND/OR substantial contributions to acquisition analysis or interpretation of data, AND (2) drafting or revising the manuscript, AND (3) final approval of the manuscript. There is no intention to use professional writers.

## Data Availability

No datasets were generated or analysed during the current study. All relevant data from this study will be made available upon study completion.

## Acknowledgments

We thank our nursing staff Alma D. Leon, R.N. and Judith Krentzel, N.P. for their review of the DIT protocols.

## References

1. Center for Disease Control and Prevention. National Diabetes Statistics Report website. [Internet]. [cited 2023 Jul 12]. Available from: https://www.cdc.gov/diabetes/data/statistics-report/index.html

2. Yang W, Dall TM, Beronjia K, Lin J, Semilla AP, Chakrabarti R, et al. Economic Costs of Diabetes in the U.S in 2017. Diabetes Care 2018. 2018 Mar 22;41(5):917–28.

3. Abdul-Ghani MA, DeFronzo RA. Mitochondrial dysfunction, insulin resistance, and type 2 diabetes mellitus. Curr Diab Rep. 2008 Jun;8(3):173–8.

4. Butler AE, Janson J, Bonner-Weir S, Ritzel R, Rizza RA, Butler PC. Beta-Cell Deficit and Increased ␤-Cell Apoptosis in. 2003;52.

5. Jay Widmer R, Lerman A. Endothelial dysfunction and cardiovascular disease. Glob Cardiol Sci Pract. 2014 Oct;2014(3):43.

6. Venables MC, Jeukendrup AE. Physical inactivity and obesity: links with insulin resistance and type 2 diabetes mellitus. Diabetes Metab Res Rev. 2009 Sep;25(S1):S18–23.

7. Kirwan JP, Sacks J, Nieuwoudt S. The essential role of exercise in the management of type 2 diabetes. Cleve Clin J Med. 2017 Jul;84(7 suppl 1):S15– 21.

8. Borghouts LB, Keizer HA. Exercise and Insulin Sensitivity: A Review. Int J Sports Med. 2000 Jan;21(1):1–12.

9. Mu L, Cohen AJ, Mukamal KJ. Resistance and Aerobic Exercise Among Adults With Diabetes in the U.S. Diabetes Care. 2014 Aug 1;37(8):e175–6.

10. Korkiakangas EE, Alahuhta MA, Husman PM, Keinänen-Kiukaanniemi S, Taanila AM, Laitinen JH. Motivators and barriers to exercise among adults with a high risk of type 2 diabetes - a qualitative study: Motivators and barriers to exercise among adults. Scand J Caring Sci. 2011 Mar;25(1):62–9.

11. Kelly S, Martin S, Kuhn I, Cowan A, Brayne C, Lafortune L. Barriers and Facilitators to the Uptake and Maintenance of Healthy Behaviours by People at Mid-Life: A Rapid Systematic Review. Wang Y, editor. PLOS ONE. 2016 Jan 27;11(1):e0145074.

12. Vranish JR, Bailey EF. Inspiratory Muscle Training Improves Sleep and Mitigates Cardiovascular Dysfunction in Obstructive Sleep Apnea. Sleep. 2016 Jun 1;39(6):1179–85.

13. Craighead DH, Tavoian D, Freeberg KA, Mazzone JL, Vranish JR, DeLucia CM, et al. A multi-trial, retrospective analysis of the antihypertensive effects of high-resistance, low-volume inspiratory muscle strength training. J Appl Physiol. 2022 Oct 1;133(4):1001–10.

14. Craighead DH, Heinbockel TC, Freeberg KA, Rossman MJ, Jackman RA, Jankowski LR, et al. Time-Efficient Inspiratory Muscle Strength Training Lowers Blood Pressure and Improves Endothelial Function, NO Bioavailability, and Oxidative Stress in Midlife/Older Adults With Above-Normal Blood Pressure. J Am Heart Assoc. 2021 Jul 6;10(13):e020980.

15. Leon BM. Diabetes and cardiovascular disease: Epidemiology, biological mechanisms, treatment recommendations and future research. World J Diabetes. 2015;6(13):1246.

16. Ramos-Barrera GE, DeLucia CM, Bailey EF. Inspiratory muscle strength training lowers blood pressure and sympathetic activity in older adults with OSA: a randomized controlled pilot trial. J Appl Physiol. 2020 Sep 1;129(3):449–58.

17. Fornoni A, Raij L. Metabolic syndrome and endothelial dysfunction. Curr Hypertens Rep. 2005 Mar;7(2):88–95.

18. Sansbury BE, Hill BG. Regulation of obesity and insulin resistance by nitric oxide. Free Radic Biol Med. 2014 Aug;73:383–99.

19. Bahadoran Z, Mirmiran P, Ghasemi A. Role of Nitric Oxide in Insulin Secretion and Glucose Metabolism. Trends Endocrinol Metab. 2020 Feb;31(2):118–30.

20. Hartge MM, Unger T, Kintscher U. The endothelium and vascular inflammation in diabetes. Diab Vasc Dis Res. 2007 Jun;4(2):84–8.

21. Suzuki T, Hirata K, Elkind MSV, Jin Z, Rundek T, Miyake Y, et al. Metabolic syndrome, endothelial dysfunction, and risk of cardiovascular events: The Northern Manhattan Study (NOMAS). Am Heart J. 2008 Aug;156(2):405–10.

22. Tavoian D, Ramos-Barrera LE, Craighead DH, Seals DR, Bedrick EJ, Alpert JS, et al. Six Months of Inspiratory Muscle Training to Lower Blood Pressure and Improve Endothelial Function in Middle-Aged and Older Adults With Above-Normal Blood Pressure and Obstructive Sleep Apnea: Protocol for the CHART Clinical Trial. Front Cardiovasc Med. 2021 Nov 24;8:760203.

23. Inaba Y, Chen JA, Bergmann SR. Prediction of future cardiovascular outcomes by flow-mediated vasodilatation of brachial artery: a meta-analysis. Int J Cardiovasc Imaging. 2010 Aug;26(6):631–40.

24. Matthews DR, Hosker JR, Rudenski AS, Naylor BA, Treacher DF, Turner RC, et al. Homeostasis model assessment: insulin resistance and fl-cell function from fasting plasma glucose and insulin concentrations in man.

25. Celermajer DS, Sorensen KE, Gooch VM, Spiegelhalter DJ, Miller OI, Sullivan ID, et al. Non-invasive detection of endothelial dysfunction in children and adults at risk of atherosclerosis. The Lancet. 1992 Nov;340(8828):1111–5.

26. Corretti MC, Anderson TJ, Benjamin EJ, Celermajer D, Charbonneau F, Creager MA, et al. Guidelines for the Ultrasound Assessment of Endothelial-Dependent Flow-Mediated Vasodilation of the Brachial Artery. J Am Coll Cardiol. 2002 Jan;39(2):257–65.

27. Harney DJ, Hutchison AT, Hatchwell L, Humphrey SJ, James DE, Hocking S, et al. Proteomic Analysis of Human Plasma during Intermittent Fasting. J Proteome Res. 2019 May 3;18(5):2228–40.

28. Kruse R, Krantz J, Barker N, Coletta RL, Rafikov R, Luo M, et al. Characterization of the CLASP2 Protein Interaction Network Identifies SOGA1 as a Microtubule-Associated Protein. Mol Cell Proteomics. 2017 Oct;16(10):1718–35.

29. James J, Valuparampil Varghese M, Vasilyev M, Langlais PR, Tofovic SP, Rafikova O, et al. Complex III Inhibition-Induced Pulmonary Hypertension Affects the Mitochondrial Proteomic Landscape. Int J Mol Sci. 2020 Aug 8;21(16):5683.

30. Sherman BT, Hao M, Qiu J, Jiao X, Baseler MW, Lane HC, et al. DAVID: a web server for functional enrichment analysis and functional annotation of gene lists (2021 update). Nucleic Acids Res. 2022 Jul 5;50(W1):W216–21.

31. Huang DW, Sherman BT, Lempicki RA. Systematic and integrative analysis of large gene lists using DAVID bioinformatics resources. Nat Protoc. 2009 Jan;4(1):44–57.

32. DeLucia CM, De Asis RM, Bailey EF. Daily inspiratory muscle training lowers blood pressure and vascular resistance in healthy men and women. Exp Physiol. 2018 Feb 1;103(2):201–11.

33. Corrêa APS, Figueira FR, Umpierre D, Casali KR, Schaan BD. Inspiratory muscle loading: a new approach for lowering glucose levels and glucose variability in patients with Type 2 diabetes. Diabet Med. 2015 Sep;32(9):1255–7.

34. Pinto MB, Bock PM, Schein ASO, Portes J, Monteiro RB, Schaan BD. Inspiratory Muscle Training on Glucose Control in Diabetes: A Randomized Clinical Trial. Int J Sports Nutr Exerc Metab. 2020 Nov 27;31(1):21–31.

